# Associations between maternal complications during pregnancy and childhood asthma: a retrospective cohort study in southern China

**DOI:** 10.1101/2022.03.21.22272680

**Authors:** Jie Tang, Ying Ma, Yu Wu, Ting Jiao, Shuangshuang Guo, Dongying Zhang, Jiewen Yang, Nali Deng, Zhijiang Liang, Harry H.X. Wang, Wei Bao, Xiaoqin Liu

## Abstract

**Background:** The associations between maternal complications during pregnancy and childhood asthma have rarely been investigated in low and middle-income countries. We aimed to investigate the associations among the southern Chinese population, and to determine whether the associations were mediated through preterm birth, cesarean delivery, low birth weight and non-breasting feeding in the first 6 months or modified by child’s lifestyles (smoking, body mass index(BMI), and sleep duration).

**Methods:** We conducted a retrospective cohort study of 208,190 children in Guangzhou, China. Information on maternal gestational hypertension, gestational diabetes, gestational anemia, and hepatitis B during pregnancy was extracted from medical records. Ever diagnosis of asthma of the children was obtained by questionnaire. We used binomial logistic regression models to estimate the adjusted odds ratios (aORs) and 95% confidential interval (CI) for childhood asthma. We conducted mediation analyses to estimate the indirect effects of gestational complications on childhood asthma mediated through preterm birth, cesarean delivery, low birth weight, and non-breastfeeding in the first 6 months.

**Findings:** The overall prevalence of ever diagnosed asthma was 1.3%. Gestational hypertension, gestational diabetes, gestational anemia, or hepatitis B during pregnancy was positively associated with ever diagnosed childhood asthma, with the aOR of 1.66(95%CI 1.31-2.10), 1.68(95%CI 1.40-2.02), 1.69(95%CI 1.49-1.91) and 1.54(95%CI 1.13-2.08), respectively. A stronger association was observed for 2 or more gestational complications (aOR= 2.34, 95%CI 1.71-3.23) than 1 gestational complication (aOR=1.80, 95%CI 1.62-1.99). The associations between maternal complications and childhood asthma did not differ by child’s smoking, BMI, or sleep duration, except that the aOR for maternal gestational hypertension was significantly higher in children who were active smoker (aOR=5.68, 95%CI 2.09-15.42) than those who were non-smoker (aOR=1.56, 95%CI 1.21-2.00). A small proportion of the associations were mediated through preterm birth, cesarean delivery, low birth weight, and non-breastfeeding in the first 6 months.

**Interpretation:** Gestational hypertension, diabetes, anemia, or hepatitis B during pregnancy was significantly associated with childhood asthma in the southern Chinese population, and the associations were partially explained by the mediation effects of cesarean delivery, preterm birth, low birthweight and non-breastfeeding in the first 6 months.

**Author summary:** *Why was this study done?:*

1. Early stage of life is a sensitive period for the development of respiratory health, but there is rarely study investigated the associations between maternal complications during pregnancy and childhood asthma in low and middle income countries.
2. Birth outcomes, breastdfeeding and children’s lifestyles are associated with athma, but it remain unclear whether the effects of maternal complications during the pregnancy on childhood asthma could be moderated by lifestyles of children or mediated through birth outcomes and breastfeeding.

*What did the researchers do and find?:*

1. In this large cohort study of more than 220,000 participants, we found gestational hypertension, diabetes, anemia, or hepatitis B during pregnancy was significantly associated with childhood ever diagnosed asthma.
2. A stronger association was observed for 2 or more gestational complications than 1 gestational complication.
3. The associations between maternal complications and childhood asthma did not differ by child’s smoking, body mass index, or sleep duration, but a small proportion of the associations were mediated through preterm birth, cesarean delivery, low birth weight, and non-breastfeeding in the first 6 months.

*What do these findings mean?:*

1. The findings suggest that interventions to address childhood asthma should consider potential associations with maternal complications.
2. To avoid cesarean delivery, preterm birth, low birthweight and non-breastfeeding in the first 6 months may reduce the effects of maternal complication during pregnancy on childhood asthma.

## 1. Introduction

Childhood asthma is a public health concern worldwide, with a prevalence of 1.8-27.4% in children aged 6-7 years and 2.6-30.5% in children aged 13-14 years, varying across countries and between sex[1]. Although mortality of childhood asthma is relatively low, childhood asthma represents a significant burden on both the family and society and ranks among the top 20 conditions worldwide for disability-adjusted life years in children[2]. Therefore, it is crucially important to understand the risk factors of childhood asthma.

Early stage of life is a sensitive period for the development of respiratory health[3], and several studies examined the associations between maternal complications during pregnancy and asthma risk in the offspring[4-14]. However, findings from these studies were mixed: some studies suggested exposure to maternal preeclampsia was associated with an increased risk of asthma in the offspring[4, 6], while others suggested a null association[5, 7, 9, 14]. Additionally, current knowledge of the associations between maternal complications and childhood asthma is predominantly derived from high-income countries[4-12], which may not be generalizable to middle and low-income countries with different epidemiological characteristics of gestational complications. For instance, pregnant women in China have a higher prevalence of maternal anemia and hepatitis B than in high-income countries[15, 16]. Moreover, the effects of maternal complications during the pregnancy on childhood asthma may be mediated through birth outcomes and feeding models [5] or modified by child’s lifestyles [17-19], which have rarely been investigated so far.

In light of these considerations, we carried out a retrospective cohort study in southern China to investigate the associations of gestational hypertension, gestational diabetes, gestational anemia, and hepatitis B during pregnancy with asthma among children aged 6 to 14 years, and whether the associations were mediated through birth outcomes and breasting feeding or modified by child’s lifestyles (smoking, body mass index (BMI), and sleep duration). In China, these four maternal complications are common and require routine examinations during pregnancy[15, 16]. The routine examinations for these four complications have covered all pregnant women in some cities and were well recorded[20], which provided baseline information for our study.

## 2 Methods

### 2.1 Data sources

This study was embedded within the framework of the Physical Health Monitoring Project of Primary and High School Students in Guangzhou (PHMPPHSS), launched by the education bureau of Guangzhou since 2017. The PHMPPHSS aimed to promote physical health and reduce common diseases of primary and junior high school students through physical examination, health education, health promotion, and policy making, which is one of the top ten livelihood events of Guangzhou Municipal Government[21]. Project-related design, organization, and implementation have been described previously[22]. Parents whose child planned to participate in the annual physical examination were contacted through the internet to get parental consent to grant permission to their children to participate in the PHMPPHSS. A pre-developed health management software was shared with the parents after they provided the written informed consent. The parents then registered in the health management software, in which a structured parental questionnaire and a child’s questionnaire were designed to collect data.

The parental questionnaire included the profile of the index child (age, sex, number of siblings, residential region, medical history, including asthma diagnosis); mothers’ age, educational level, and smoking during pregnancy, family monthly income; and feeding models of the index child in the first 6 months. Parental questionnaire also included pre-pregnancy body weight and height of mother; delivery patterns; gestational weeks; complications during pregnancy, and the birth weight of the index child, all of which were asked to answer based on medical records during pregnancy and the index child’s birth certificate.

The child’s questionnaire collected information on active smoking [23], and duration of sleep[24], answered by the child with the help of their parents. Duration of sleep was evaluated by items derived from the Pittsburgh Sleep Quality Index. Completeness of parental and child’s questionnaires was checked by the physicians in each school upon submission. The data was cleaned, compiled, and de-identified by Health Promotion Centre for Primary and Secondary Schools of the Education Bureau of Guangzhou. Trained qualified health workers did physical examinations and collected blood samples for the children. They measured body weight and height using calibrated instruments.

### 2.2 Study population

This study is reported as per the Strengthening the Reporting of Observational Studies in Epidemiology (STROBE) guideline (S1 Checklist). We conducted a retrospective cohort study in 233,507 students aged 6 to14 years who participated in PHMPPHSS from September 2017 to June 2018 in 446 primary schools and 155 junior high schools across 11 administrative districts of Guangzhou. We excluded 9,064 children whose mothers did not answer the parental questionnaire; 63 children whose mothers had pre-existing hypertension or type 1 or 2 diabetes; 6,063 children whose mothers did not respond to gestational complications; 8,678 children whose mothers did not respond to asthma diagnosis of the index child. We further excluded 1,201 children who were multiple live births and 248 children who had missing data on age or sex. The remaining 208,190 children were included in the final analysis. Supplementary figure S1 shows the selection of participants for the present study.

The PHMPPHSS was approved by the Institutional Review Board of the Education Bureau of Guangzhou. Written informed consent was obtained from the participants’ parents before enrolling in this study. The present study was executed jointly by Guangzhou Education Bureau and Guangzhou Medical University. The review boards waived the ethical approval due to the usage of de-identified data.

### 2.3 Exposure definition and ascertainment

Children were categorized as exposed if their mothers reported the diagnosis of or met the diagnostic criteria of any of the following four complications during pregnancy: gestational hypertension (defined as blood pressure greater than or equal to 140 mmHg systolic or 90 mmHg diastolic on two separate occasions at least four hours apart after 20 weeks of pregnancy when previous blood pressure was normal[25]), gestational diabetes (defined as any degree of hyperglycaemia for the first time during pregnancy[26]), gestational anemia (defined as hemoglobin (Hb) <11 g/dL, or hematocrit <33%, at any time during the pregnancy[27]), and hepatitis B (defined as HBsAg is positive regardless of the status of HBeAg, at any time during the pregnancy[28]). Mothers who reported having or met the diagnostic criteria of the above-mentioned gestational complications were asked to provide medical records through health management software, for ascertaining. Mothers who reported having gestational complication(s) without providing medical records were interviewed by the physician through telephone, recording the detailed information about gestational complication(s) reported, such as when and where the gestational complication(s) was diagnosed and what the main symptoms were. Health Promotion Centre for Primary and Secondary Schools of the Guangzhou Education Bureau was responsible for external quality assessment. One hundred mothers who reported not having gestational complications were randomly selected in each administrative district and required to provide medical records. Interclass correlation coefficients expressing between-person variance, obtained by analysis of replicate pairs of these mothers from all administrative districts involved, were all higher than 0.99.

#### Outcome

The outcome was physician-diagnosed asthma (ever asthma) in the children. Information on asthma diagnosis was obtained by the parental questionnaire based on the International Study on Asthma and Allergy in Childhood (ISAAC) Questionnaire (“Has your child ever had asthma diagnosis by a doctor?”)[29], which was also ascertained by the physicians through on-site consultation, physical examinations, reviewing medical records and previous related auxiliary examinations. This validated approach has been used in several nationwide epidemiological studies of asthma in China[30].

### 2.4 Statistical analysis

The data analysis was performed from 1^st^ March 2021 to 1^st^ March 2023. Mean and standard deviation (SD) were calculated for age. Frequencies and proportions were used to describe the characteristics of the children and their mothers. Two-tailed, unpaired *t* or *χ*^*2*^ tests were used to compare the distribution of the outcome according to different characteristics of the children and their mothers.

Binomial logistic regression models were employed to estimate the adjusted odds ratios (aORs) and 95% confidential interval (CI) for childhood asthma by gestational hypertension, gestational diabetes, gestational anemia, and hepatitis B, respectively, and two stages of adjustment were used to examine the robustness of the associations. We selected potential confounding factors which are known to be associated with maternal complications and the outcomes. In model 1, we adjusted for children’s sex (boy or girl), age (continuous variable), residential region (urban or rural), one-child family (yes or no), maternal age (younger than 20 years; 21-35 years; or older than 35 years), mother’s educational level (junior high school or below; senior high school; or college or above) and family monthly income (<8,000 RMB; 8000-15000 RMB; or >15000RMB). In model 2, we additionally adjusted for potential confounders, including mothers’ BMI before pregnancy (underweight [BMI<18.5]; healthy weight [18.5≤BMI<24]; overweight or obesity [BMI≥24]), mutually adjusted for gestational complications, and maternal smoking during pregnancy (yes or no). We also examined the association between the number of maternal gestational complications and the offspring’s asthma to determine whether multiple maternal complications during pregnancy have a synergistic effect on childhood asthma [31]. Because there were 0.06% mothers had 3 or 4 gestational complications, we categorized the number of gestational complications into three groups: 0, 1, and 2 or more.

We further conducted subgroup analyses to examine whether the associations between maternal complications during pregnancy and childhood asthma were modified by the children’s lifestyles, including smoking (yes or no), BMI (underweight/healthy weight or overweight/obesity), sleep duration (adequate or inadequate). BMI was calculated by dividing the weight in kg by the square of height in m. Childrens’ BMI z-score was calculated according to Chinese standard and was categorized into underweight/healthy weight (BMI z-score <1) and overweight/obesity (BMI z-score ≥1)[32]. Duration of sleep was classified into adequate or inadequate sleep according to the National Sleep Foundation’s recommendation[33]. Because the prevalence of childhood asthma varied with age and residential region, we also examined whether the association between maternal complications during pregnancy and childhood asthma varied with child’s age (6∼12 years, or 13∼14 years) and residential region (urban and rural). Among all the subgroup analyses, we adjusted for the covariates as we did in model 2. Differences within subgroup analyses were assessed by the test for interactions of pregnancy complications and each lifestyle factor.

Previous studies suggested that cesarean delivery, preterm birth, low birthweight and non-breastfeeding in the first 6 months could mediate the associations between maternal complications and childhood asthma [5, 7, 8], we therefore conducted mediation analyses by a logistic decomposition of the total effects into direct and indirect effects using the “ldecomp” command in the Stata[34]. We also estimated the indirect effects of these potential mediators separately. Figure 1 shows the theoretical framework underlying our mediation analysis.

**Figure 1.**
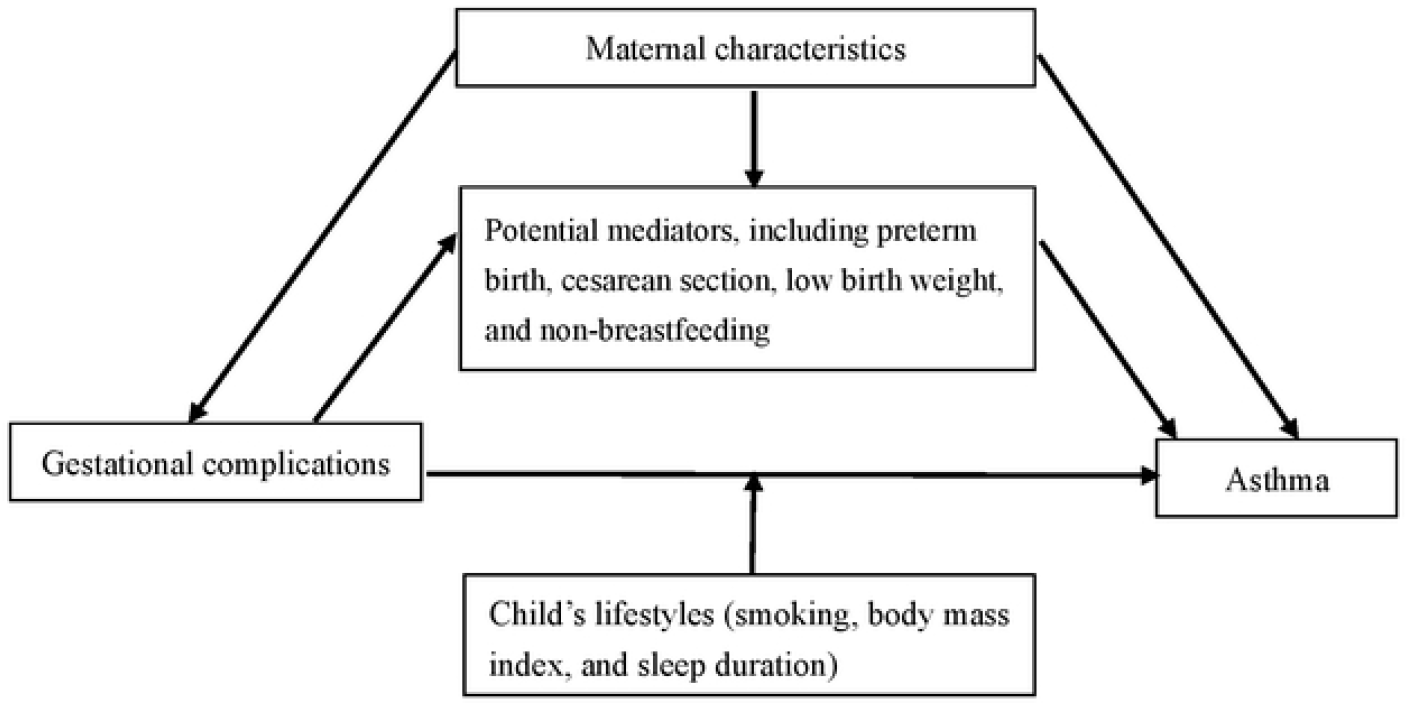
Directed acyclic graph used to guide thw analyses.

Data were missing in family monthly income (8623, 4.1%), maternal pre-pregnancy BMI (6247, 3.0%), children’s BMI (8516, 4.1%) and smoking (1441, 0.7%). We imputed these missing covariates by using the monotone logistic regression method based on other sociodemographic covariates by creating 20 imputed datasets[35]. The significance level was set at *P*=0.05 and all tests were 2-sided. Statistical analyses were conducted by Stata (version 14.0, StataCorp LLC, College Station, TX), Statistic software R (version 3.6.2, https://www.R-project.org/)..

## 3. Results

### 3.1 Population description

Of 208,190 children included in this study, 44.0% were from five urban administrative districts of Guangzhou City, 80.7% were primary school students and 54.2% were boys. The sample size and the proportion of primary school students in each administrative district are shown in supplementary Table S1. The age of children ranged from 6 to 14 years, with a mean age of 9.5 (SD=2.5) years. Overall, 2,696 (1.3%, 95%CI 1.2%-1.4%) children had ever been diagnosed with asthma. Compared to children without asthma, children with asthma were more likely to live in urban areas, be boys, older, born through cesarean delivery, preterm birth, and overweight/obesity, and have inadequate sleep duration (Table 1).

**Table 1.** Characteristics of children and their mothers.

| Variables | Ever Asthma |  | Total<br>(N=208,190) | P |
| --- | --- | --- | --- | --- |
|  | No (N=205,494) | Yes(N=2,696) |  |  |
| Children’s characteristics |  |  |  |  |
| Residential area (n, %) |  |  |  | <0.001 |
| Unban | 90,103(43.9) | 1,502(55.7) | 91,605(44.0) |  |
| Suburb | 115,391(56.1) | 1,194(44.3) | 116,585(56.0) |  |
| Sex (n, %) |  |  |  | <0.001 |
| Boys | 110,979(54.0) | 1,848(68.6) | 112,827(54.2) |  |
| Girls | 94,515(46.0) | 848(31.4) | 95,363(45.8) |  |
| Age, y (Mean, SD) | 9.5(2.5) | 9.7(2.4) | 9.5(2.5) | <0.001 |
| Single child (n, %) | 91,882(55.3) | 1,461(54.2) | 115,073(55.3) | 0.256 |
| Cesarean delivery (n, %) | 74,372(36.2) | 1,345(49.9) | 75,672(36.4) | <0.001 |
| Preterm birth (n, %) | 14,117(6.9) | 308(11.4) | 14,425(6.9) | <0.001 |
| Low birthweight (n, %) | 7,434(3.6) | 122(4.5) | 7,556(3.6) | 0.012 |
| Non-breastfeeding (n, %) | 127,804(62.2) | 1,724(64.0) | 129,528(62.2) | 0.062 |
| **Participants' BMI <sup>a</sup>** |  |  |  |  |
| Underweight/Health weight | 163,737(83.1) | 2,042(79.1) | 165,779(83.1) | <0.001 |
| Overweight/Obesity | 33,357(16.9) | 538(20.9) | 33,895(17.0) |  |
| **Adequate sleep (n, %)** | 103,916(50.6) | 1,182(43.8) | 105,098(50.1) | <0.001 |
| **Active smoking (n, %) <sup>a</sup>** | 4,419 (2.2) | 61(2.3) | 4,480(2.2) | 0.687 |
| **Mothers' characteristics** |  |  |  |  |
| **Maternal age (Mean, SD)** | 27.7(4.9) | 27.5(4.9) | 27.7(4.9) | 0.071 |
| **Educational level (n, %)** |  |  |  | <0.001 |
| Junior high school or below | 14,454(7.0) | 170(6.3) | 14,624(7.0) |  |
| Senior high school | 100,259(48.8) | 1,135(42.1) | 101,394(48.7) |  |
| College or above | 90,781(44.2) | 1,391(51.6) | 92,172(44.3) |  |
| **Smoking during pregnancy (n, %)** | 1,505(0.7) | 41(1.5) | 1,546(0.7) | <0.001 |
| **Family monthly income (n, %) <sup>a</sup>** |  |  |  | <0.001 |
| <8,000 RMB | 89,799(43.7) | 1,125(41.7) | 90,924(43.7) |  |
| 8000~15,000 RMB | 70,581(34.3) | 828(30.7) | 71,409(34.3) |  |
| >15,000 RMB | 36,550(17.8) | 684(25.4) | 37,234(17.9) |  |
| **Maternal pre-pregnancy BMI <sup>a</sup>** |  |  |  |  |
| Underweight (BMI<18.5) | 51,458(25.8) | 420(15.6) | 51,878(25.7) | <0.001 |
| Healthy weight (18.5≤BMI<24) | 134,584(67.5) | 2046(76.2) | 136,630(67.7) |  |
| Overweight/Obesity (≥24) | 13,217(6.6%) | 218(8.1) | 13,435(6.7) |  |

<sup>a</sup> missing data existed

### 3.2 Association between maternal complications during pregnancy and childhood asthma

A total of 2,722(1.3%), 4,364(2.1%), 13,789 (6.6%), and 1,915(0.9%) of children were born to mothers who had gestational hypertension, gestational diabetes, gestational anemia or hepatitis B during pregnancy, respectively. The prevalence of asthma among children born to mothers with gestational hypertension was 2.8%; gestational diabetes, 3.0%; gestational anemia, 2.1%; and hepatitis B, 2.4%. Gestational hypertension, gestational diabetes, gestational anemia or Hepatitis B during pregnancy was positively associated with childhood asthma. The aORs are presented in Table 2. The aORs reduced substantially from model 1 to model 2 except for gestational anemia. In the fully adjusted model (model 2 in Table 2), the aOR for gestational hypertension was 1.66 (95%CI, 1.31-2.10); gestational diabetes, 1.68 (95%CI, 1.40-2.02); gestational anemia, 1.69 (95%CI, 1.49-1.91); and hepatitis B, 1.54 (95%CI, 1.13-2.08).

**Table 2.**
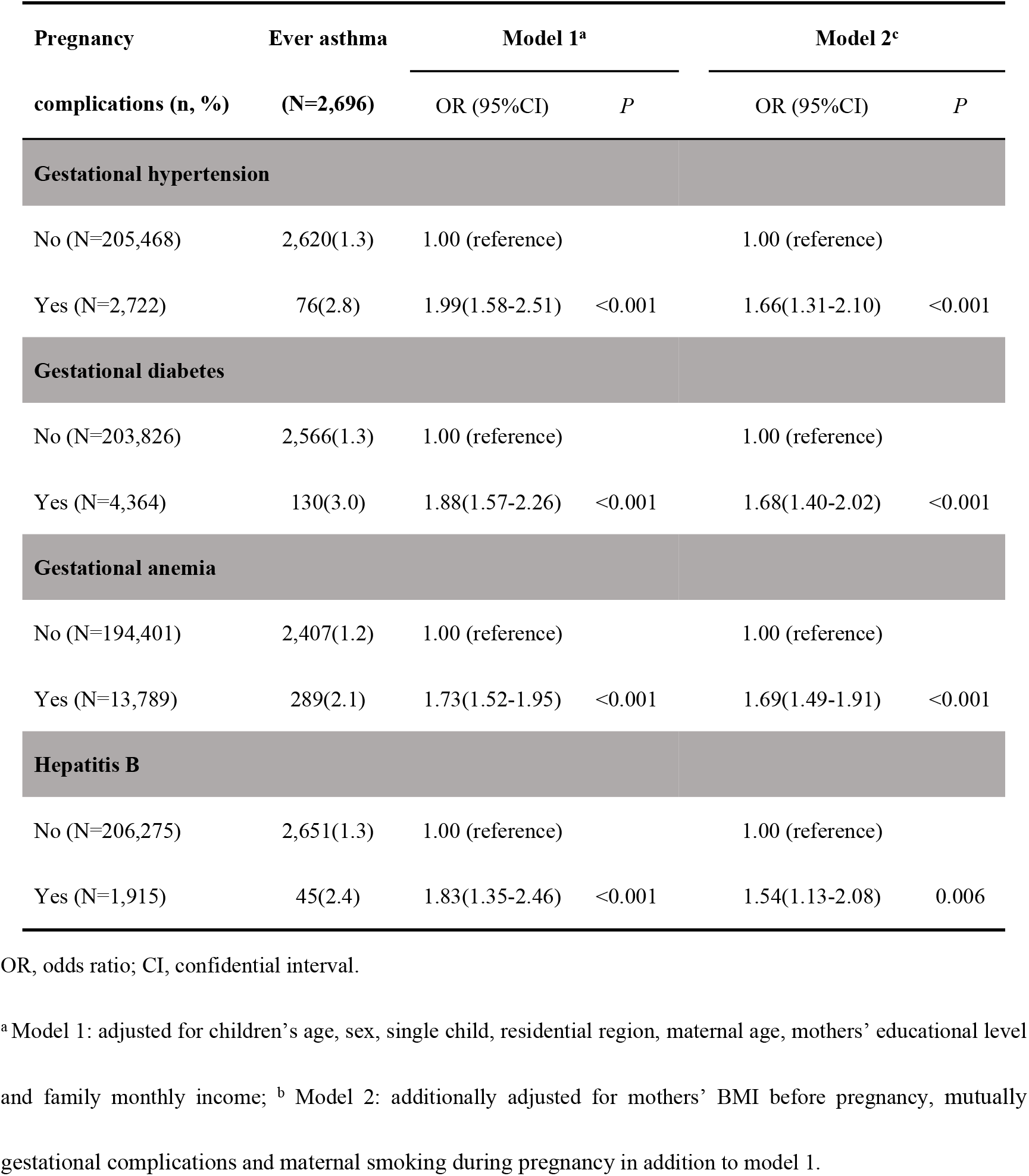
Associations between maternal gestational complications and childhood asthma.

| Pregnancy | Ever asthma | Model 1<sup>a</sup> |  | Model 2<sup>c</sup> |  |
| --- | --- | --- | --- | --- | --- |
| complications (n, %) | (N=2,696) | OR (95%CI) | *P* | OR (95%CI) | *P* |
| Gestational hypertension |  |  |  |  |  |
| No (N=205,468) | 2,620(1.3) | 1.00 (reference) |  | 1.00 (reference) |  |
| Yes (N=2,722) | 76(2.8) | 1.99(1.58-2.51) | <0.001 | 1.66(1.31-2.10) | <0.001 |
| Gestational diabetes |  |  |  |  |  |
| No (N=203,826) | 2,566(1.3) | 1.00 (reference) |  | 1.00 (reference) |  |
| Yes (N=4,364) | 130(3.0) | 1.88(1.57-2.26) | <0.001 | 1.68(1.40-2.02) | <0.001 |
| Gestational anemia |  |  |  |  |  |
| No (N=194,401) | 2,407(1.2) | 1.00 (reference) |  | 1.00 (reference) |  |
| Yes (N=13,789) | 289(2.1) | 1.73(1.52-1.95) | <0.001 | 1.69(1.49-1.91) | <0.001 |
| Hepatitis B |  |  |  |  |  |
| No (N=206,275) | 2,651(1.3) | 1.00 (reference) |  | 1.00 (reference) |  |
| Yes (N=1,915) | 45(2.4) | 1.83(1.35-2.46) | <0.001 | 1.54(1.13-2.08) | 0.006 |

OR, odds ratio; CI, confidential interval.
<sup>a</sup> Model 1: adjusted for children's age, sex, single child, residential region, maternal age, mothers' educational level and family monthly income; <sup>b</sup> Model 2: additionally adjusted for mothers' BMI before pregnancy, mutually gestational complications and maternal smoking during pregnancy in addition to model 1.

### 3.3 Association between number of maternal complications during pregnancy and childhood asthma

Altogether, 20,039 (9.6%,) and 1,285 (0.6%) mothers reported to have 1, and 2 or more gestational complication(s) during pregnancy. The prevalence of asthma among children who had exposed to 1, and 2 or more gestational complication (s) was 2.3% and 3.1%, respectively; while the prevalence of asthma among children who did not expose to any gestational complication was 1.2%. The aORs are presented in Table 3. Both the aORs did not change substantially from model 1 to model 3. In the fully adjusted model (model 2 in Table 3), the aOR for 1 gestational complication was 1.80 (95%CI, 1.62-1.99), and an increased aOR was observed for 2 or more gestational complications (2.34, 95%CI, 1.71-3.23).

**Table 3.** Association between number of maternal gestational complications and childhood asthma a.

| Number of gestational<br>complications (n, %) | Ever asthma<br>(N=2,696) | Model 1<sup>b</sup> |  | Model 2<sup>d</sup> |  |
| --- | --- | --- | --- | --- | --- |
|  |  | OR (95%CI) | *P* | OR (95%CI) | *P* |
| 0 (N=186,866) | 2,205 (1.2) | 1.00 (reference) |  | 1.00 (reference) |  |
| 1 (N=20,039) | 451(2.3) | 1.82(1.64-2.02) | <0.001 | 1.80(1.62-1.99) | <0.001 |
| ≥2 (N=1,285) | 40(3.1) | 2.41(1.75-3.32) | <0.001 | 2.34(1.71-3.23) | <0.001 |

OR, odds ratio; CI, confidential interval.
<sup>a</sup> maternal gestational complications include gestational hypertension, gestational diabetes, gestational anemia and hepatitis B.
<sup>b</sup> Model 1: adjusted for children's age, sex, single child, residential region, maternal age, mothers' educational level and family monthly income; <sup>c</sup> Model 2: adjusted for mothers' BMI and maternal smoking during pregnancy in addition to model 1.

### 3.3 Subgroup analysis

The associations of gestational hypertension (Figure 2-A), gestatational diabetes (Figure 2-B), gestational anemia (Figure 2-C) and hepatitis B (Figure 2-D) with childhood asthma did not appear to be modified by child’s smoking status, BMI category and sleep duration. However, the aOR for asthma for gestational hypertension was significantly higher in children who were active smoker (5.68, 95%CI 2.09-15.42) than non-smokers (1.56, 95%CI 1.21-2.00) (Figure 2-A). The associations were also similar according to child’s age group, and residential region (See in supplementary Table S2 and S3).

**Figure 2.**
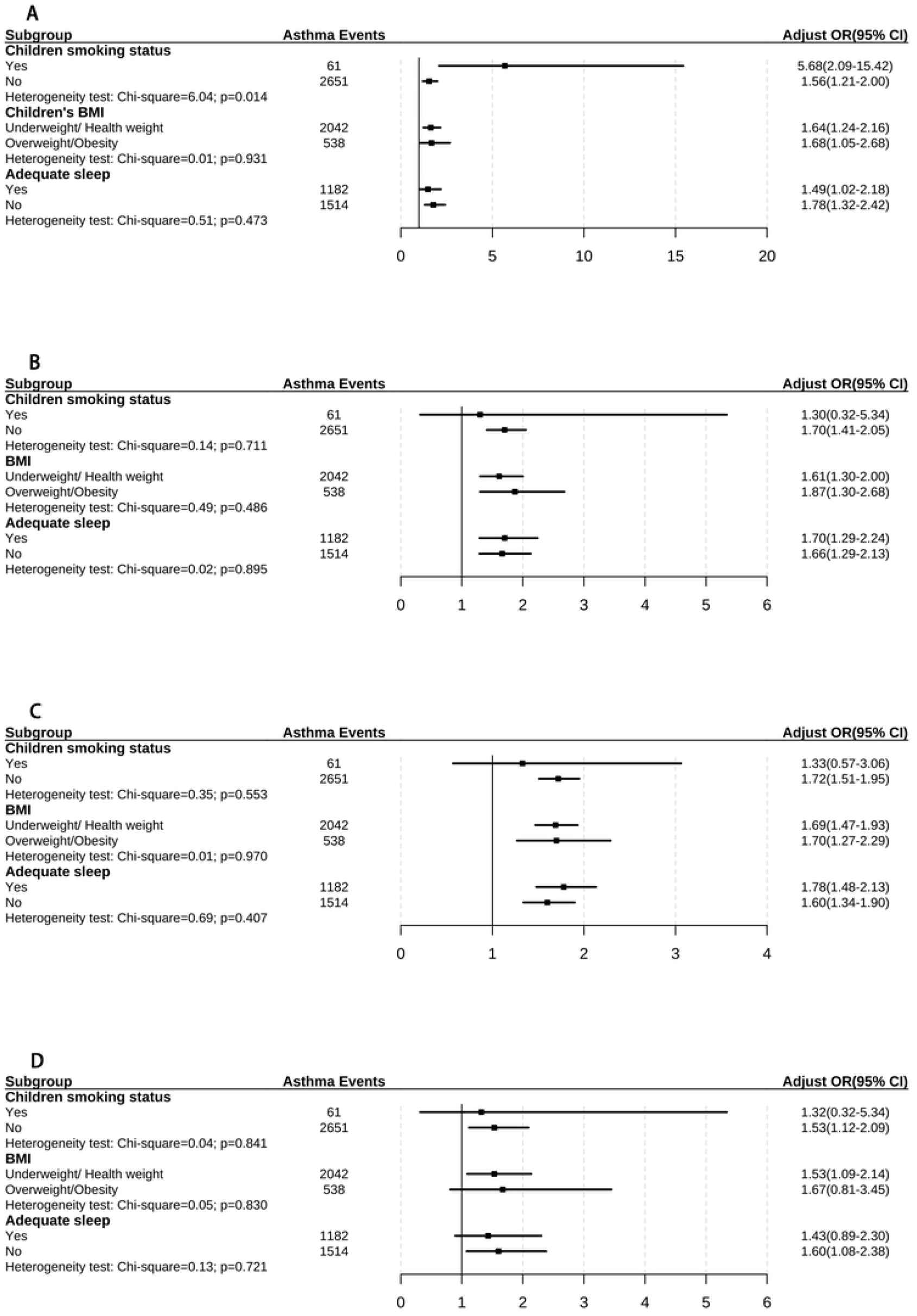
Subgroup analyses of associations between maternal complications and chilhood asthma.

### 3.4 Mediation analysis

The results from the mediation analyses indicated a significant direct effect of gestational hypertension (aOR=1.43, 95%CI, 1.13-1.80), gestational diabetes (aOR=1.65, 95%CI 1.36-1.97), gestational anemia (aOR=1.67, 95%CI 1.46-1.90), or hepatitis B (aOR=1.54, 95%CI 1.15-2.05) on childhood asthma. The combined indirect effects of cesarean delivery, preterm birth, low birthweight and non-breastfeeding in the first 6 months on associations between gestational complications and childhood asthma were also significant, the aOR was 1.21 (95%CI 1.16-1.26) for gestational hypertension, 1.14(95%CI 1.11-1.17) for gestational diabetes, 1.02 (95%CI 1.01-1.03) for gestational anemia and 1.04 (95%CI 1.03-1.05) for hepatitis B (Table 4).

**Table 4.**
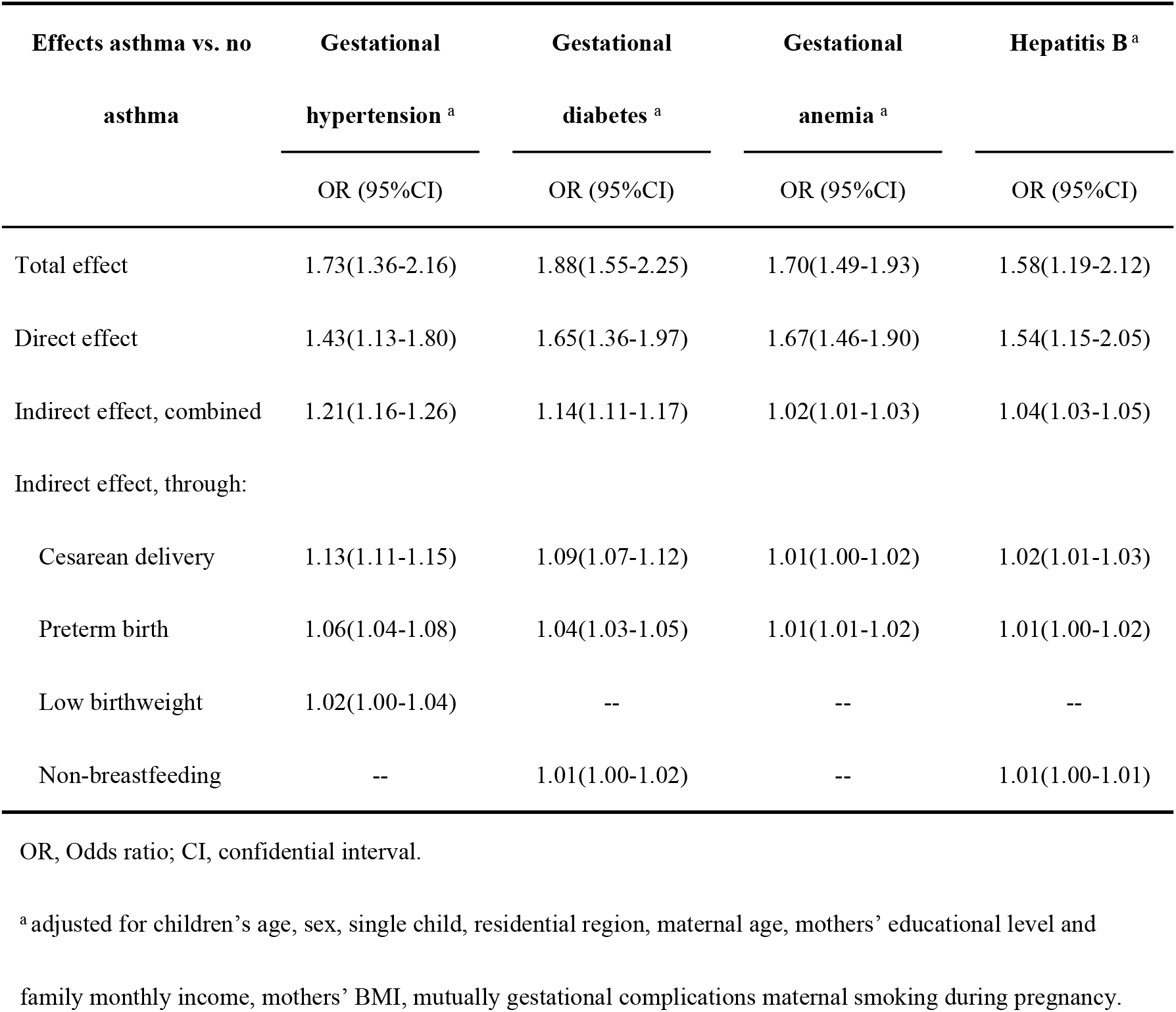
Logistic regression model of direct and indirect effects through mediators of gestational complications on childhood asthma

## Discussion

In this study among the southern Chinese population, we found that maternal gestational hypertension, gestational diabetes, gestational anemia, or hepatitis B was associated with childhood asthma, respectively. A stronger association was observed for 2 or more gestational complications than 1 gestational complication. The associations were not modified by children’s smoking, BMI, sleep duration and child’s age group, except that the aOR for maternal gestational hypertension was significantly higher in children who were active smoker than those who were non-smoker. A small proportion of the associations were mediated through cesarean delivery, preterm birth, low birthweight and non-breastfeeding in the first 6 months.

### Comparison with previous studies

Several studies have examined the association between gestational hypertension and childhood asthma, but the results were inconsistent[4-7]. Two studies from Denmark reported an increased risk of asthma among children born to mothers with preeclampsia, although the magnitude of the relative risks differed slightly[4, 6]. While a study based on Avon Longitudinal Study found that gestational hypertension or preeclampsia was not associated with asthma at 7 years old[7].

Studies that examined the association between gestational diabetes and childhood asthma also demonstrated inconsistent results[8, 10, 13]. Martinez and colleagues[8] found that the risk of childhood asthma was increased for gestational diabetes requiring medication (hazard ratio[HR] =1.12, 95%CI 1.01-1.25) but not for gestational diabetes not requiring medication (HR=1.01, 95%CI 0.93-1.10). Liu and colleagues[13] found that gestational diabetes was associated with small increased risk of early-onset transient wheezing (prevalence ratio[PR]=1.08, 95%CI 1.00-1.17) and early-onset persistent wheezing (PR=1.15, 95%CI 1.05-1.26). However, a pooled analysis of 14 European birth cohorts found a null association between diabetes and the risk of wheezing up to 24 months of the children[10].

Four studies examined the association between gestational anemia and childhood asthma, and the results were also mixed[9, 11, 12, 14]. Nafstad et al. [9] found that exposure to gestational anemia was associated with an increased risk of childhood asthma (OR=1.67, 95%CI 1.24-2.36). Triche and colleagues[11] found that gestational anemia was associated with recurrent infant wheeze in the first year (OR=2.17, 95%CI 1.18-4.00), and wheezing before 3 years (OR=2.42, 95%CI 3.18-4.23). While the third study based on the Generation R study and the fourth study based on Avon Longitudinal Study suggested that gestational anemia was not associated with childhood asthma[12, 14].

The discrepancies on the associations of gestational hypertension, gestational diabetes, or gestational anemia with childhood asthma among previous studies may be related to the different prevalences of these gestational complications, different measurements and criteria of childhood asthma, study design, adjustment strategies, and sample size with small studies failing to find an association. Our findings that gestational hypertension, gestational diabetes, or gestational anemia during pregnancy was independently associated with childhood asthma are consistent with most of the previous studies conducted in high-income countries[4-6, 8, 9, 11, 13], although the prevalence of childhood asthma is lower than that in high-income countries[1].

### Explanations of maternal complications associated with childhood asthma

Potential biological mechanisms linking maternal pregnancy complications and childhood asthma remain unknown but may involve multiple pathways. The intrauterine environment provides the substrate for many important processes, including lung and immune system development, and support of optimal fetal growth requires adequate maternal nutritional status[11]. Gestational hypertension, gestational diabetes, gestation anemia, and hepatitis B during pregnancy may predispose the fetuses to stress, chronic inflammation, hypoxia, malnutrition status, and fetal hyperinsulinemia, which in turn may lead to immune suppression and interfere with lung growth and maturation; alveolar sac formation, proliferation, and expansion; and impaired surfactant production which then predispose offspring to newborn respiratory conditions and chronic lung disease such as asthma[8, 36, 37]. Our study and a previous study suggested that the association between maternal complications during pregnancy and childhood asthma may partially be due to intermediate factors, such as preterm birth, cesarean delivery, and low birth weight[5], therefore, mechanisms linking these intermediate factors and childhood asthma could also be explanations of maternal complications associated with childhood asthma. Of noted that findings in previous study suggested the association between maternal complications during pregnancy and childhood asthma may be largely due to intermediate factors[5], is different from our findings, which need further study.

### Strengths and limitations of this study

It is the first study that evaluated the independent associations between four common maternal gestational complications and childhood asthma in a Chinese population. The large sample size allowed us to examine whether the associations between maternal gestational complications and childhood asthma were modified by child’s lifestyles. Mediation analyses in this study allowed us to estimate the direct effects not be mediated through intermediate variables. Furthermore, it is also the first study that investigated the association of Hepatitis B and number of maternal complications with childhood asthma, which filled in the knowledge gap of the association between maternal complications during pregnancy and childhood asthma.

Some important limitations should be acknowledged. Firstly, information bias may exist since data were collected by self-reported questionnaires in this study. Mothers may recall their gestational complications incorrectly, and mothers with pre-existing or/and diabetes may be diagnosed during pregnancy initially, both of which would lead to misclassification of exposure. However, the prevalences of the four maternal gestational complications and childhood asthma are comparable with the prevalences reported previously[38-41]. Moreover, our quality assessment shows significant interclass correlations. Together, these suggest that the information bias is minimal. Secondly, we did not have data on phenotypes of asthma, we can not examine whether the associations with exposures of interest are specific for different asthma phenotypes. Thirdly, we do not have data on the severity of complications and treatment, which hampers us from analyzing the associations between severity of complications and childhood asthma. Fourthly, although we have adjusted for many important confounding factors, potential confounding bias owing to unmeasured factors such as family history of asthma, air pollution, allergies, or genetic susceptibility could not be ruled out. Fifthly, although our population is representative of Guangzhou residents, these results may not be generalizable to other populations with different population characteristics. Finally, causal inferences should be cautious since this is an observational study.

## Conclusions

In the southern Chinese population, we found that gestational hypertension, gestational diabetes, gestational anemia, or hepatitis B are significantly associated with childhood asthma, and a stronger association was observed for 2 or more gestational complications than 1 gestational complication. The associations between maternal gestational complications and childhood asthma were not modified by child’s lifestyles, but could be partially mediated through cesarean delivery, preterm birth, low birthweight and non-breastfeeding in the first 6 months.

## Data Availability

Data generated and/or analyzed during the current study can be shared publicly by contacting the corresponding author (Professor Jie Tang). All data on the PHMPPHSS are available from the Health Promotion Centre for Primary and Secondary Schools, the Education Bureau of Guangzhou (Miss Nali Deng) on reasonable request for researchers.

## Acknowledgment

We would like to thank the participants and physicians in each participated school for their valuable contributions.

## Contributors

JT, YM, YW contributed to the conception and design, acquisition of data, analyses and interpretation of the data, drafted the article, revised it critically for important intellectual content and gave final approval of the version to be published. TJ, SG, DZ, JW, ND, ZL, HHXW, WB and XL contributed to the conception and design, acquisition of data, revised the drafted manuscript critically for important intellectual content and gave final approval of the version to be published.

